# Faster reaction times of CSF Alpha-Synuclein Seed Amplification Assay predict the Diffuse Malignant subtype of Parkinson’s Disease at 10-year follow-up

**DOI:** 10.1101/2025.03.27.25324778

**Authors:** Piergiorgio Grillo, Giulietta Maria Riboldi, Antonio Pisani, Un Jung Kang, Seyed-Mohammad Fereshtehnejad

**Author notes:** Corresponding authors **Un Jung Kang, MD** NYU Grossman School of Medicine, 435 E 30th Street, SB1013, New York, NY 10016, USA, 1-212-263-8179’ **S.M. Fereshtehnejad, MD PhD,** The Edmond J. Safra Program in Parkinson’s Disease and the Morton and Gloria Shulman Movement Disorders Clinic, Toronto Western Hospital, 7MCL-402, 399 Bathurst Street Toronto, Ontario, Canada M5T 2S8.

## Abstract

**Background:** Data-driven approaches identified Mild Motor Predominant (MMP), Intermediate (IM), and Diffuse Malignant (DM) as Parkinson’s Disease (PD) subtypes with different motor and non-motor impairment at diagnosis. It remains unclear whether these subtypes remain stable over time or whether they represent distinct biological substrates. The alpha-synuclein seed amplification assay in CSF (CSF-αSyn-SAA) might provide further insights.

**Objective:** to evaluate the association between baseline CSF-αSyn-SAA parameters and 10-year clinical evolution of PD subtypes.

**Methods:** 323 sporadic PD patients from PPMI dataset were classified as MMP, IM, or DM at baseline and 10-year follow-up based on motor, cognitive, sleep and dysautonomia features. CSF-αSyn-SAA parameters were collected at baseline using a 150-hrs protocol. CSF Aβ1-42, tTau and pTau181, CSF and serum NfL were also considered at baseline.

**Results:** Reaction times (T50, TTT) and area under the curve (AUC) respectively were shorter and larger in DM compared to IM/MMP. The difference in baseline amplification parameters was more evident when comparing subtypes based on 10-year clinical features (T50, η2=0.036; TTT, η2=0.031; AUC, η2=0.033; all p values < 0.05) than when comparing subtypes based on baseline clinical features (T50, η2=0.012; TTT, η2=0.012; AUC, η2=0.013; all p<0.05). Shorter T50 and TTT at baseline were associated with greater risk of DM versus MMP at 10-year follow-up (T50, OR=3.3, 95%CI: 1.3-8.1, p=0.010; TTT, OR=4.6, 95%CI: 1.8-11.6, p=0.001). Aβ, Tau and NfL were similar between groups.

**Conclusions:** Baseline CSF-αSyn-SAA parameters predicted long-term PD progression. Faster reactions were associated with a more severe 10-year PD phenotype considering motor and non-motor features.

**Plain Language Summary:** *Background:* The diagnosis of Parkison’s Disease is drastically changing by the development of Alpha-Synuclein Seed Amplification Assay. The assay enables, for the first time, the detection of pathological forms of alpha-synuclein in cerebrospinal fluid in living patients. Alpha-Synuclein Seed Amplification is very accurate in discerning individuals with Parkinson’s Disease versus healthy subjects, but it remains unknown whether it can also inform about prognosis.

*Objective:* We assessed the ability of the assay to predict the 10-year clinical progression of Parkinson’s Disease.

*Methods:* Public data from an international cohort were used. At time of diagnosis, we classified 323 individuals with Parkinson’s Disease into three clinical subtypes: Mild Motor Predominant, Intermediate, and Diffuse Malignant. These subtypes were characterized by a progressively increasing burden of motor and non-motor symptoms. Subjects with available follow-up data were re-classified using the same subtypes after 10 years of disease. Time-dependent signal changes of Alpha-Synuclein Seed Amplification Assay in Cerebrospinal Fluid were measured at baseline and used to predict the 10-year phenotype.

*Results:* Firstly, we found that subtypes were not stable categories. Around a half of participants changed subtype over time, mostly shifting towards a more aggressive one. Notably, our results showed that faster reactions on Alpha-Synuclein Seed Amplification Assay at baseline were associated with a 10-year phenotype more aggressive in terms of motor symptoms, dysautonomia, sleep and cognitive impairment, i.e the Diffuse Malignant subtype.

*Conclusion:* Characteristics of the assay underlying the final positivity or negativity outcomes performed at a milder and early stage of PD may identify a subgroup of subjects that are more likely to undergo a more rapid clinical deterioration. Further studies, however, are needed to confirm and expand this result.

## INTRODUCTION

Phenotypic heterogeneity is a well-recognized trait of Parkinson’s Disease (PD)(1,2). The identification of PD subtypes is important for prognostic implication and understanding pathophysiological basis that could lead to more rationale treatment. Subtypes have been initially based on the motor phenotypes (3,4). More recently data driven approaches and cluster analysis allowed to identify subtypes without a priori hypothesis (5–8). Recent works by Fereshtehnejad et al was able to identify three main PD phenotypes, namely a Mild Motor Predominant (MMP), an Intermediate (IM), and a Diffuse Malignant (DM), leveraging unsupervised cluster analysis of different motor and non-motor symptoms (REM sleep behavior disorder (RBD), dysautonomia, early cognitive impairment) in large clinical cohorts (7,8). Subtypes have been noted to change over time within individual subjects, but most studies have been limited to relatively short periods of no longer than 3-4 years(9,10). Whether these subtypes represent a snapshot of between-subject variability or biological differences that persist over time is unclear. The lack of imaging or biochemical biomarkers has prevented establishing the connections between clinical presentation and underlying biology.

Seed amplification assays (SAA), originally developed for prions disease, has been welcomed as the first reliable *in vivo* biomarker for PD diagnosis(11–13). Beyond the binary outcome (positive/negative), the parameters of the amplification kinetics of alpha-synuclein SAA on CSF (CSF-αSyn-SAA) may provide information about the α-syn conformation and properties of aggregates(12,14,15). A few studies demonstrated that amplification parameters correlate with certain clinical features (e.g., dysautonomia and cognitive impairment) (15–18). Recently, the speed of seeding was shown to predict the phenoconversion of idiopathic RBD (iRBD) to PD (19). However, it remains unknown whether CSF-αSyn-SAA parameters at baseline may serve as a biomarker that can predict long-term clinical progression of PD.

Here, we classified 323 PD subjects from Parkinson’s Progression Markers Initiative (PPMI) repository into MMP, IM and DM at baseline and after 10 years of follow-up. The clinical trajectory of these subtypes over time was assessed. CSF-αSyn-SAA amplification parameters at the time of diagnosis were compared in MMP versus IM/DM and used to predict the long-term phenotype. Markers of axonal degeneration (i.e Neurofilament Light Chain) and Alzheimer’s disease (AD) co-pathology (i.e. Amyloid Beta Peptide, Total and Serine 181 Phosphorylated Tau) were also considered for a comprehensive analysis.

## METHODS

### Participants

Clinical and biochemical data were obtained from the PPMI cohort (data-cut: 20240729). PPMI is an international multi-center study collecting clinical, genetical, radiological, and biochemical markers from subjects with PD longitudinally followed-up. As per PPMI design, diagnosis of PD is based on abnormal dopamine transporter (DAT)-SPECT and two of either resting tremor, bradykinesia, rigidity, or asymmetric resting tremor, or asymmetric bradykinesia. Only subjects with a sporadic form of disease and positivity on the CSF-αSyn-SAA (n=323) were considered for this study. A description of the CSF-αSyn-SAA is provided below. PD participants carrying pathogenic variants in any of the following genes - *GBA, LRRK2, PINK1, VPS35* and *PRKN* - were excluded.

### Clinical Assessment

Demographic information and clinical features were collected at baseline (n=323) and 10 years follow-up when available (n=146). Baseline assessment was performed within 2 years from diagnosis in a drug-naïve condition. The following parameters were considered for the participants: sex, age, disease duration, Movement Disorder Society-Unified PD Rating scale (MDS-UPDRS) part III in off state, part II and IV, Hoehn and Yahr scale (H&Y) in off state, postural instability– gait difficulty (PIGD) score, levodopa equivalent daily dose (LEDD), RBD Screening Questionnaire (RBDSQ), Scales for Outcomes in PD-Autonomic dysfunction (SCOPA-AUT), Montreal cognitive assessment (MoCA), Benton judgment of line orientation (BJLO), Hopkins Verbal Learning Test (HVLT), Letter number sequencing (LNS), Semantic fluency test (SF), Symbol digit test (SDT).

### Clinical Subtyping

Each subject was assigned to either Mild Motor Predominant (MMP), Intermediate (IM) or Diffuse Malignant (DM) PD subtype based on composite scores including motor and non-motor components (RBD, dysautonomia, early cognitive impairment)(7). The motor component was calculated with MDS-UPDRS part II, III and PIGD scores. The non-motor components included three domains: domain 1 (RBDSQ), domain 2 (SCOPA-AUT), domain 3 (BJLO, HVLT, LNS, SF, SDT). MMP was defined as motor and all three non-motor scores below the 75th percentile calculated on the baseline cohort (n=323); DM as either motor score plus ≥ 1/3 non-motor score > 75th percentile, or all three non-motor scores > 75th percentile; subjects not included in the previous categories were labelled as IM. The subtype membership was calculated at baseline for all participants (n=323) by means of clinical scores obtained in proximity of diagnosis, at a drug-naïve stage. A smaller subgroup (n=146) had long-term follow-up information which were used to re-define the clinical subtype after 10 years of disease. In the latter case, new percentiles, based on the 10-year follow-up evaluation, were used to adjust the classification for the clinical worsening that each participant inevitably experienced during the years. Baseline and 10-year follow-up subtypes for a single subject could coincide or not (subtype shifting) depending on the specific disease course.

### Biochemical Assessment: Alpha-Synuclein Seed Amplification Assay

Technical aspects of the CSF-αSyn-SAA were discussed in a previous paper(12). CSF-αSyn-SAA was run in triplicates. The reaction lasted 150 hours and provided a curve with the following parameters: Fmax (highest raw fluorescence from each well; RFU), T50 (time to reach 50% of the Fmax; hours), TTT (time to reach a 5,000 RFU threshold; hours), Slope (RFU/hours) and AUC (area under the curve; RFU*hours). A categorical outcome (positive/negative/inconclusive) was based on the fluorescence of three replicates. Only PD subjects with a positivity on CSF-αSyn-SAA were included in the study (n=323). A negative (n=23 subjects) or inconclusive (n=4 subjects) response was excluded. The mean of three replicates was calculated for each amplification parameter. The CSF-αSyn-SAA was performed exclusively at baseline.

### Biochemical Assessment: AD Biomarkers and Neurofilament Light Chain

CSF AD biomarkers including Amyloid Beta Peptide 1-42 (Aβ1-42), Total Tau (tTau), Phosphorylated Tau at Serine 181 (pTau181), were considered for the study in addition to the ratios of tTau/Aβ1-42 and pTau181/Aβ1-42. CSF and Serum NfL levels were also analyzed. Elecsys (Roche) and Simoa-Quanterix (Biogen) assays were used for dosages on CSF and Serum respectively. All the above measurements referred to the baseline assessment and were available for the following subsample of subjects: CSF AD biomarkers for 311, CSF NfL for 169, and Serum NfL for 284 out of 323 participants.

### Statistical analysis

Distribution of variables was evaluated with Shapiro-Wilk test. Categorical variables were compared by Chi-Square test followed by Post-Hoc Test with Bonferroni-Adjustment. Comparison between groups was assessed by Kruskal-Wallis’s Test; Dunn-Bonferroni Post-Hoc Analysis for multiple comparisons was applied; eta squared (η2) was used to measure the effect size. For parametric variables one-way ANOVA and Bonferroni Post-Hoc Analysis were used. Multinomial Logistic Regression (MLR) was used to predict the 10-year PD clinical subtype given baseline CSF-αSyn-SAA parameters. Only CSF-αSyn-SAA parameters whose mean/distribution significantly differed between the 10-year PD clinical subtypes were included in the model.

Multicollinearity was defined by Variation Inflation Factor (VIF) greater than 5.0. Predictors were expressed as quantiles above and below the median. Significance was set at p<0.05. Statistical analysis was performed by using IBM-SPSS Version-28.

### Standard Protocol Approvals, Registrations, and Patient Consents

Information about ethical approval and patient consents can be retrieved on the PPMI website (www.ppmi-info.org).

### Data Availability

Data are available from authors upon reasonable request.

## RESULTS

### Clinical Signature of Mild Motor Predominant, Intermediate and Diffuse Malignant PD subtypes

Following the assignment rules described above, MMP subtype was characterized by milder motor and non-motor disability compared to the IM/DM both at baseline and 10-year follow-up (Table 1). At baseline, MMP scored lower on MDS-UPDRS-part III in OFF state, HY in OFF state, SCOPA-AUT and RBDSQ (p<0.001 for all listed variables), with no sex, age, race, ethnicity, years of education and disease duration differences between groups (Table 1). Similar results were found at 10-year follow-up between MMP vs IM/DM (MDS-UPDRS-part III in OFF state, p=0.002; HY in OFF state, p<0.001; SCOPA-AUT, p<0.001; RBDSQ, p<0.001) (Table 1). The mean (±SD) LEDD at 10-year follow-up across subtypes was 915.1 (±673.7) without significant differences between subtypes.

**Table 1.** Comparison of clinical-demographic features between Mild Motor Predominant, Intermediate and Diffuse Malignant subtypes of Parkinson’s Disease at baseline (n=323) and 10-year follow-up (n=146). Values were given in mean (±standard deviation). Kruskal-Wallis/Chi-Square p value: statistical significance is marked in bold. Post-Hoc p value: *p<0.005; **p<0.001. Abbreviations: MMP, Mild Motor Predominant; IM, Intermediate; DM, Diffuse Malignant; n, number; y, years; m, male; f, female; LEDD, levodopa equivalent daily dose; MDS-UPDRS-part III, Movement Disorder Society-Unified PD Rating scale part III; MDS-UPDRS-part IV, Movement Disorder Society-Unified PD Rating scale part IV; HY, Hoehn and Yahr scale; MoCA, Montreal cognitive assessment; SCOPA-AUT, Scales for Outcomes in PD-Autonomic dysfunction; RBDSQ, RBD Screening Questionnaire.

|  | **Mild Motor**<br>**Predominant** | **Intermediate** | **Diffuse Malignant** | **p value** | **Post Hoc**<br>**Analysis** |
| --- | --- | --- | --- | --- | --- |
| **Baseline** |  |  |  |  |  |
| n | 182 | 101 | 40 |  |  |
| Sex (f, %) | 66, 36.2% | 37, 36.6% | 12, 30% | p=0.730 | - |
| Age (y) | 61.1 (9.3) | 62.5 (9.2) | 63.6 (9.9) | p=0.180 | - |
| Race (White, %) | 162, 94.2% | 90, 90.0% | 34, 89.5% | p=0.552 | - |
| Ethnicity (Hispanic, %) | 3, 1.7% | 4, 4.0 % | 0, 0.0 % | p=0.290 | - |
| Education (y) | 15.8 (2.6) | 15.3 (2.6) | 15.5 (3.3) | p=0.439 | - |
| Duration (y) | 0.6 (0.6) | 0.5 (0.5) | 0.7 (0.6) | p=0.075 | - |
| LEDD (mg/day) | 0 | 0 | 0 |  |  |
| MDS-UPDRS-part III in OFF state | 18.4 (7.5) | 22.1 (8.7) | 29.8 (9.1) | **p<0.001** | MMP versus IM*<br>MMP versus DM**<br>IM versus DM** |
| HY in OFF state | 1.5 (0.5) | 1.7 (0.5) | 1.8 (0.4) | **p<0.001** | MMP versus IM*<br>MMP versus DM** |
| MDS-UPDRS-part IV | 0 | 0 | 0 |  |  |
| MOCA | 27.4 (2.1) | 26.8 (2.5) | 26.6 (2.6) | p=0.066 | - |
| SCOPA-AUT | 6.5 (3.4) | 11.5 (6.7) | 17.1 (6.3) | **p<0.001** | All Comparisons** |
| RBDSQ | 2.8 (1.5) | 5.2 (2.8) | 7.1 (2.9) | **p<0.001** | MMP versus IM**<br>MMP versus IM**<br>IM versus DM* |
| **10-Year Follow-Up** |  |  |  |  |  |
| n | 44 | 64 | 38 |  |  |
| Sex (f, %) | 19, 43.2% | 21, 32.8% | 7, 18.4% | p=0.056 | - |
| Age (y) | 68.2<br>(8.7) | 69.0 (8.6) | 71.5 (8.3) | p=0.434 | - |
| Race (White, %) | 42, 95.5% | 61, 96.8% | 37, 97.4% | p=0.711 | - |
| Ethnicity (Hispanic, %) | 0, 0.0% | 3, 4.7% | 0, 0.0% | p=0.141 | - |
| Education (y) | 16.4 (2.7) | 15.5 (2.3) | 15.8 (2.3) | p=0.082 | - |
| LEDD (mg/day) | 841.6 (473.5) | 906.7 (592.8) | 1014.4 (950.4) | p=0.799 | - |
| MDS-UPDRS-part III in OFF state | 32.3 (12.6) | 38.7 (16.7) | 49.7 (17.8) | **p=0.002** | MMP versus DM* |
| HY in OFF state | 2.1 (0.4) | 2.1 (0.3) | 2.2 (0.5) | **p<0.001** | MMP versus DM*<br>IM versus DM* |
| MDS-UPDRS-part IV | 3.8 (3.7) | 4.1 (4.2) | 5.2 (3.6) | p=0.111 | - |
| MOCA | 27.7 (2.5) | 27.0 (2.8) | 25.4 (5.2) | p=0.368 | - |
| SCOPA-AUT | 10.4 (4.4) | 13.8 (5.7) | 20.7 (7.5) | **p<0.001** | MMP versus IM** |
| RBDSQ | 3.2 (1.9) | 5.6 (3.4) | 7.5 (3.5) | **p<0.001** | MMP versus IM*<br>MMP versus DM* |

No differences in term of sex, race, ethnicity and years education were found between baseline (n=323) and 10-year follow-up cohorts (n=146) (prevalence of female: baseline cohort, 35.6% vs 10-year follow-up cohort, 32.2%, p=0.472; prevalence of white individuals: baseline cohort, 92.3% vs 10-year follow-up cohort, 96.6%, p=0.118; prevalence of Hispanic individuals: baseline cohort, 2.3% vs 10-year follow-up cohort, 2.1%, p=0.894; years of education: baseline cohort, 15.6 vs 10-year follow-up cohort, 15.9, p=0.492).

### Stability of Clinical Subtypes Over Time

The 146 subjects with available data at BL and at 10-year follow-up were classified by the three clinical subtypes at both time points. Around half of the subjects changed subtype from baseline to follow-up (83 out of 146, 56.8%). Clinical subtype remained stable for the remainders (63 out of 146, 43.2%) (Table 2). Among subjects in the MMP subtype at baseline, 37.6% (n=35 out of 93) remained in the same group at 10-year follow-up. The rest of the subject (n=58 out of 93) moved to a less benign clinical phenotype (48.4% were classified as IM and 14.0% as DM at 10-year follow-up). In the IM at baseline, 43.9% of subjects remained within the same subtype, 19.5% were classified as MMP, and 36.6% as DM at 10-year follow-up. The DM membership at baseline was reconfirmed at 10-year follow-up for most participants (83.3%). Only two subjects changed subtype from DM to IM/MMP at follow-up.

**Table 2.** Shift of Parkinson’s Disease Subtypes from Baseline to 10-Year Follow-Up Assessment.

|  |  | Mild Motor<br>Predominant<br>At 10-Year Follow-Up | Intermediate<br>At 10-Year Follow-Up | Diffuse Malignant<br>At 10-Year Follow-Up | Total |
| --- | --- | --- | --- | --- | --- |
| Mild Motor Predominant<br>At Baseline | Count | 35 | 45 | 13 | 93 |
|  | Percentage within<br>Baseline Subtype | 37.6% | 48.4% | 14.0% | 100% |
| Intermediate<br>At Baseline | Count | 8 | 18 | 15 | 41 |
|  | Percentage within<br>Baseline Subtype | 19.5% | 43.9% | 36.6% | 100% |
| Diffuse Malignant<br>At Baseline | Count | 1 | 1 | 10 | 12 |
|  | Percentage within<br>Baseline Subtype | 8.3% | 8.3% | 83.3% | 100% |

### Amplification Parameters of CSF-αSyn-SAA and Clinical Subtypes at Baseline

Comparison between CSF-αSyn-SAA parameters and clinical subtypes at baseline showed longer T50/TTT and smaller AUC in MMP compared to IM/DM (values reported as mean±SD; T50: MMP 72.5±11.7 vs IM 69.2±9.5 vs DM 68.5±8.3 hours, χ²(2, 323) = 6.098, p=0.047; TTT: MMP 67.1±12.5 vs IM 63.6±9.1 vs DM 62.4±7.7 hours, χ²(2, 323) = 6.115, p=0.047; AUC: MMP 25792471.6±4186018.3 vs IM 26977933.6±3217124.8 vs DM 27327623.5±2792542.6 RFU*hours, χ²(2, 323) = 6.189, p=0.045) (Table 3, Figure 1). Despite an overall significance, the post-hoc analysis did not show any significant differences by the pairwise comparison.

**Figure 1.**
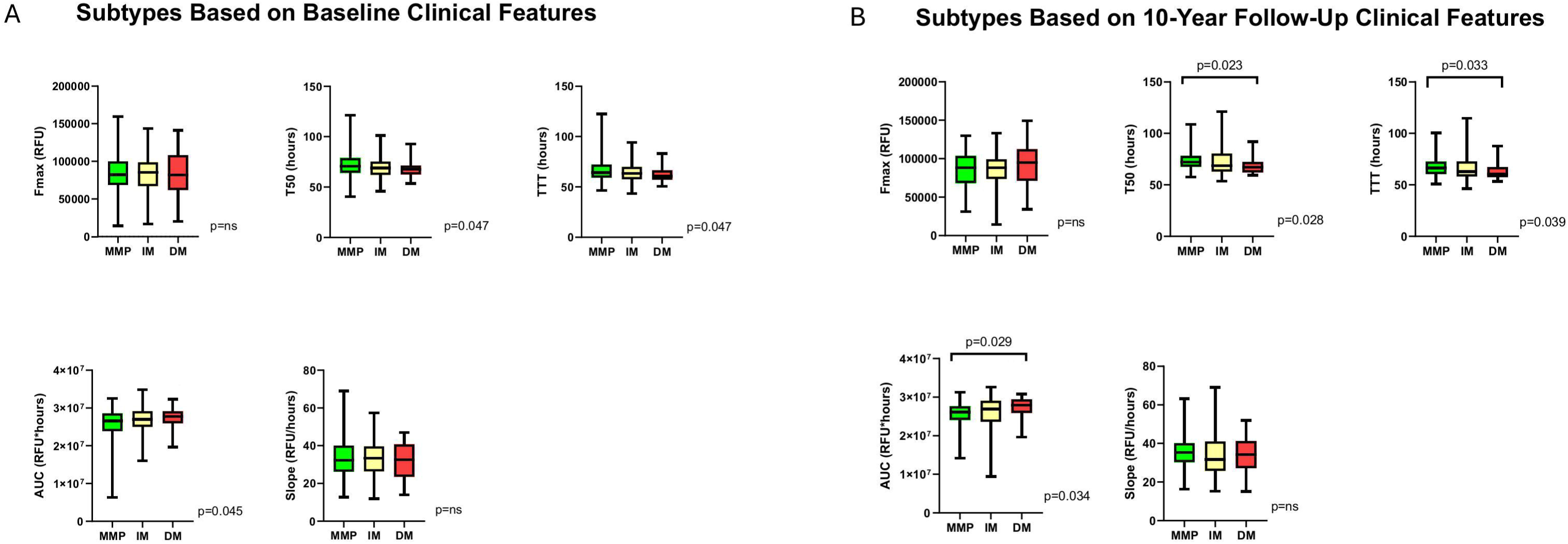
Box and whiskers plots illustrating the differences in baseline CSF-αSyn-SAA parameters across PD subtypes based on baseline (A) and 10-year follow-up (B) clinical features. Significance of ANOVA/Kruskal-Wallis’s Tests was reported in the bottom right corner of each graph. Only significant post-hoc comparisons were reported. Abbreviations: CSF-αSyn-SAA, Alpha-synuclein seed amplification assay on CSF; MMP, Mild Motor Predominant; IM, Intermediate; DM, Diffuse Malignant; T50, time to reach 50% of the Fmax; TTT, time to reach a 5,000 RFU threshold; AUC, area under the curve

**Table 3.** Comparison of CSF-αSyn-SAA amplification parameters, CSF AD biomarkers, CSF and Serum NfL between Mild Motor Predominant, Intermediate and Diffuse Malignant subtypes of Parkinson’s Disease at baseline. Values were given in mean (±standard deviation). Statistical significance is marked in bold. Abbreviations: MMP, Mild Motor Predominant; IM, Intermediate; DM, Diffuse Malignant; n, number. Fmax, highest raw fluorescence from each well; T50, time to reach 50% of the Fmax; TTT, time to reach a 5,000 RFU threshold; AUC, area under the curve; AD biomarkers, Alzheimer’s Disease biomarkers; NfL, Neurofilament Light Chain; Aβ1-42, Amyloid Beta Peptide 1-42; tTau, Total Tau; pTau181, Phosphorylated Tau at Serine 181.

|  | **Mild Motor**<br>**Predominant**<br>**at Baseline** | **Intermediate**<br>**at Baseline** | **Diffuse Malignant**<br>**at Baseline** | ***p* value** | **Post Hoc**<br>**Analysis** |
| --- | --- | --- | --- | --- | --- |
| **CSF-αSyn-SAA**<br>**Amplification Parameters at Baseline** |  |  |  |  |  |
| n | 182 | 101 | 40 |  |  |
| Fmax (RFU) | 84256.8<br>(23929.2) | 82374.0<br>(24720.8) | 85422.4<br>(29984.8) | p=0.757 | - |
| T50 (hours) | 72.5 (11.7) | 69.2 (9.5) | 68.5 (8.3) | **p=0.047** | No significant<br>comparison |
| TTT (hours) | 67.1 (12.5) | 63.6 (9.1) | 62.4 (7.7) | **p=0.047** | No significant comparison |
| AUC (RFU*hours) | 25792471.6<br>(4186018.3) | 26977933.6<br>(3217124.8) | 27327623.5<br>(2792542.6) | **p=0.045** | No significant<br>comparison |
| Slope (RFU/hours) | 33.6 (10.7) | 33.1 (9.6) | 32.0 (9.5) | p=0.922 | - |
| **CSF AD Biomarkers at Baseline** |  |  |  |  |  |
| n | 174 | 99 | 38 |  |  |
| Aβ1-42 (pg/mL) | 872.0 (328.8) | 934.0 (342.1) | 839.9 (368.9) | p=0.182 | - |
| tTau (pg/mL) | 162.6 (56.2) | 170.0 (59.0) | 176.1 (66.6) | p=0.352 | - |
| pTau181 (pg/mL) | 13.9 (5.1) | 14.4 (5.3) | 15.4 (6.5) | p=0.394 | - |
| tTau/Aβ1-42 | 0.18 (0.04) | 0.18 (0.05) | 0.24 (0.12) | p=0.327 | - |
| pTau181/Aβ1-42 | 0.015 (0.003) | 0.016 (0.004) | 0.020 (0.010) | p=0.171 | - |
| **CSF NfL at Baseline** |  |  |  |  |  |
| n | 103 | 44 | 22 |  |  |
| CSF NfL(pg/mL) | 92.2 (44.7) | 94.8 (41.9) | 104.0 (45.6) | p=0.412 | - |
| **Serum NfL at Baseline** |  |  |  |  |  |
| n | 159 | 91 | 34 |  |  |
| Serum NfL (pg/mL) | 12.2 (5.4) | 12.9 (6.5) | 14.3 (6.7) | p=0.205 | - |

### Amplification Parameters of CSF-αSyn-SAA at Baseline as Predictors of 10-Year Follow-up Clinical Subtypes

When CSF-αSyn-SAA parameters obtained at baseline were compared between the 10-year clinical subtypes, the longer T50/TTT and smaller AUC were noted in MMP compared to IM/DM (values reported as mean±SD; T50: MMP 73.9±11.2 vs IM 72.3±12.6 vs DM 68.1±7.3 hours, χ²(2, 146) = 7.150, p=0.028; TTT: MMP 68.1±11.1 vs IM 66.7±13.7 vs DM 62.0±7.4 hours, χ²(2, 146) = 6.512, p=0.039; AUC: MMP 25511406.0±3716051.8 vs IM 25834362.9±4503639.5 vs DM 27453849.4±2458992.2 RFU*hours, χ²(2, 146) = 6.744, p=0.034) (Table 4, Figure 1). At 10-year follow-up, the differences between groups were larger (10-Year Follow-Up: T50-η2=0.036; TTT-η2=0.031, AUC-η2=0.033, all p values < 0.05; Baseline: T50-η2=0.012, TTT-η2=0.012, AUC-η2=0.013, all p<0.05) and the post-hoc analysis showed a significant longer T50 and TTT, and smaller AUC at the 10-year-follow-up in MMP subtype versus DM (p=0.023; p=0.033; p=0.029). A MLR was run to predict the 10-year subtypes based on the baseline T50, TTT and AUC (Table 5, Figure 2). Amplification parameters were expressed as quantiles above and below the median. Given the presence of multicollinearity between T50, TTT and AUC, each predictor was entered separately (VIF: T50 7.55, TTT 18.41, AUC 28.04). MLR showed that subjects with T50 and TTT below the median at baseline were associated with a greater risk of being classified as DM or IM versus MMP subtype at 10-year follow-up (T50-DMvsMMP: OR=3.3, 95%CI=1.3-8.1, p=0.010; T50-IMvsMMP, OR=2.6, 95%CI=1.1-5.8, p=0.020; TTT-DMvsMMP, OR=4.6, 95%CI=1.8-11.6, p=0.001; TTT-IMvsMMP, OR=2.7, 95%CI=1.2-6.1, p=0.016) (Table 5, Figure 2). Eventually, the same three baseline amplification parameters were compared between subjects who changed subtype over time versus those who did not (Supplementary Table 1, 2, 3). Subjects with MMP subtype at baseline who shifted towards IM/DM at follow-up were associated with a higher percentage of TTT values below the median compared to subjects that remained in the MMP subtype (MMP to IM/DM vs stable MMP subtype, 51.7% vs 26.6%, p=0.029; relative risk of shifting from MMP to IM/DM if TTT below the median: OR=2.7, 95%CI=1.1-6.6, p=0.031, model fitting information - χ²(1) = 4.887, p < 0.027). A similar trend with the T50 quantile below the median was observed in subjects who shifted from IM to DM at follow-up (shifted to DM vs shifted to MMP vs stable on IM, 73.3% vs 25.0% vs 55.6%, p=0.084)

**Figure 2.**
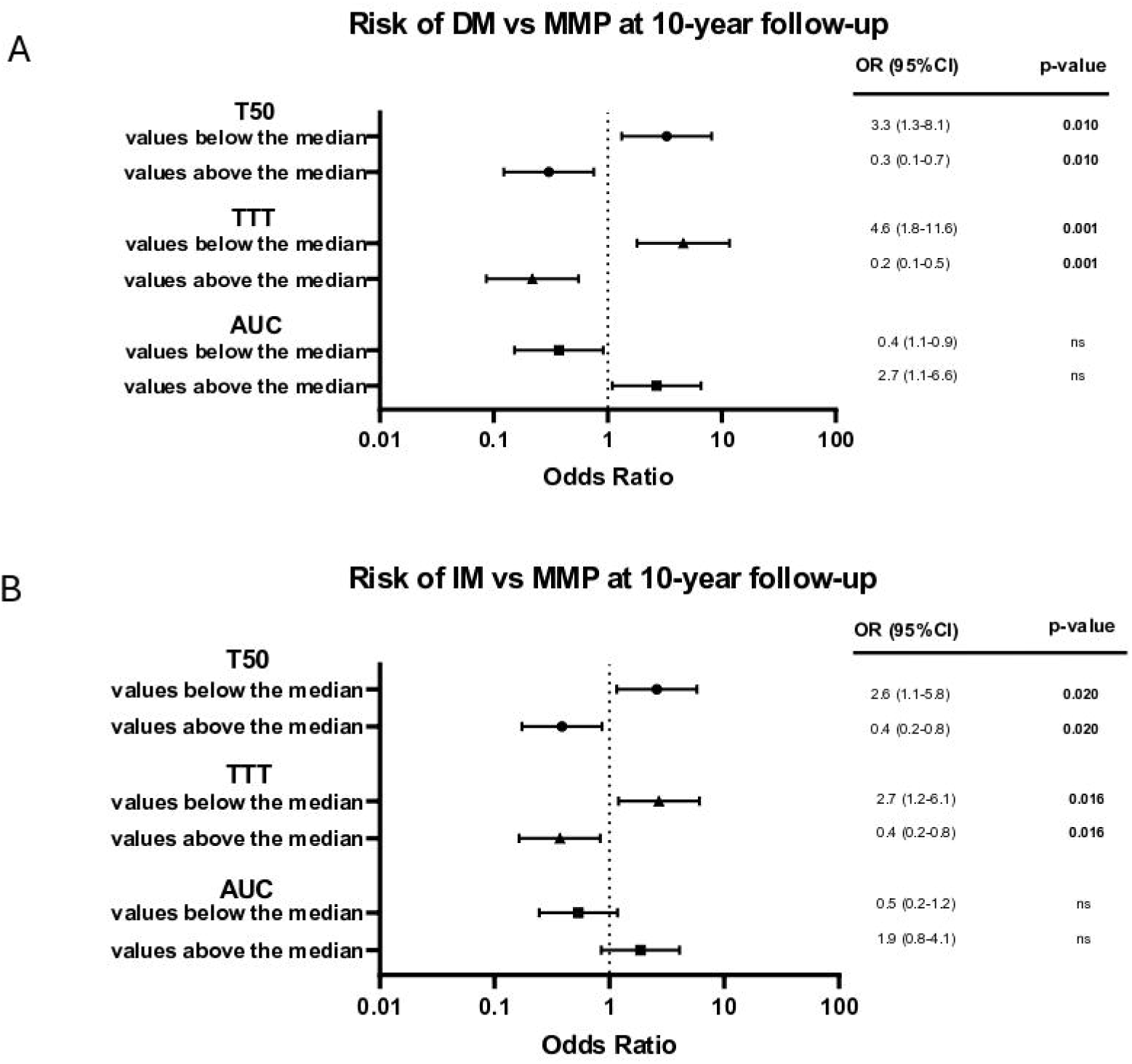
Forest plots of multinomial logistic regression comparing the odds of being classified as DM (A), or IM (B), versus MMP at 10-year follow-up, based on CSF-αSyn-SAA amplification parameters at baseline. The plots illustrate odds ratios (ORs) and 95% confidence interval (CI) for each CSF-αSyn-SAA amplification parameter, dichotomously expressed as values below and above the median. Only CSF-αSyn-SAA amplification parameters whose mean/distribution significantly differed between 10-year PD subtypes were considered. Median of T50: 69.7 hours; Median of TTT: 63.6 hours; Median of AUC: 26909171.8 RFU*hours. Statistical significance is marked in bold. Abbreviations: PD, Parkinson’s Disease; CSF-αSyn-SAA, Alpha-synuclein seed amplification assay on CSF; MMP, Mild Motor Predominant; IM, Intermediate; DM, Diffuse Malignant; T50, time to reach 50% of the Fmax; TTT, time to reach a 5,000 RFU threshold; AUC, area under the curve; OR, odds ratio, 95% CI, 95% confidence interval.

**Table 4.**
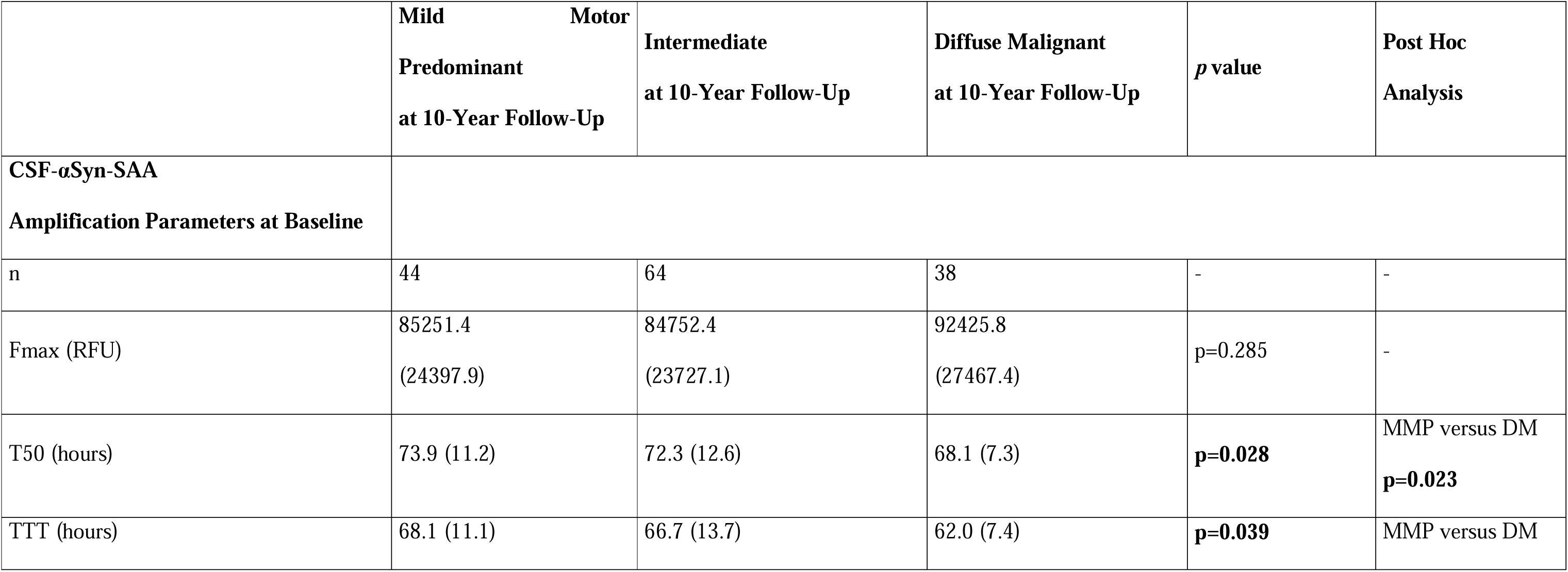
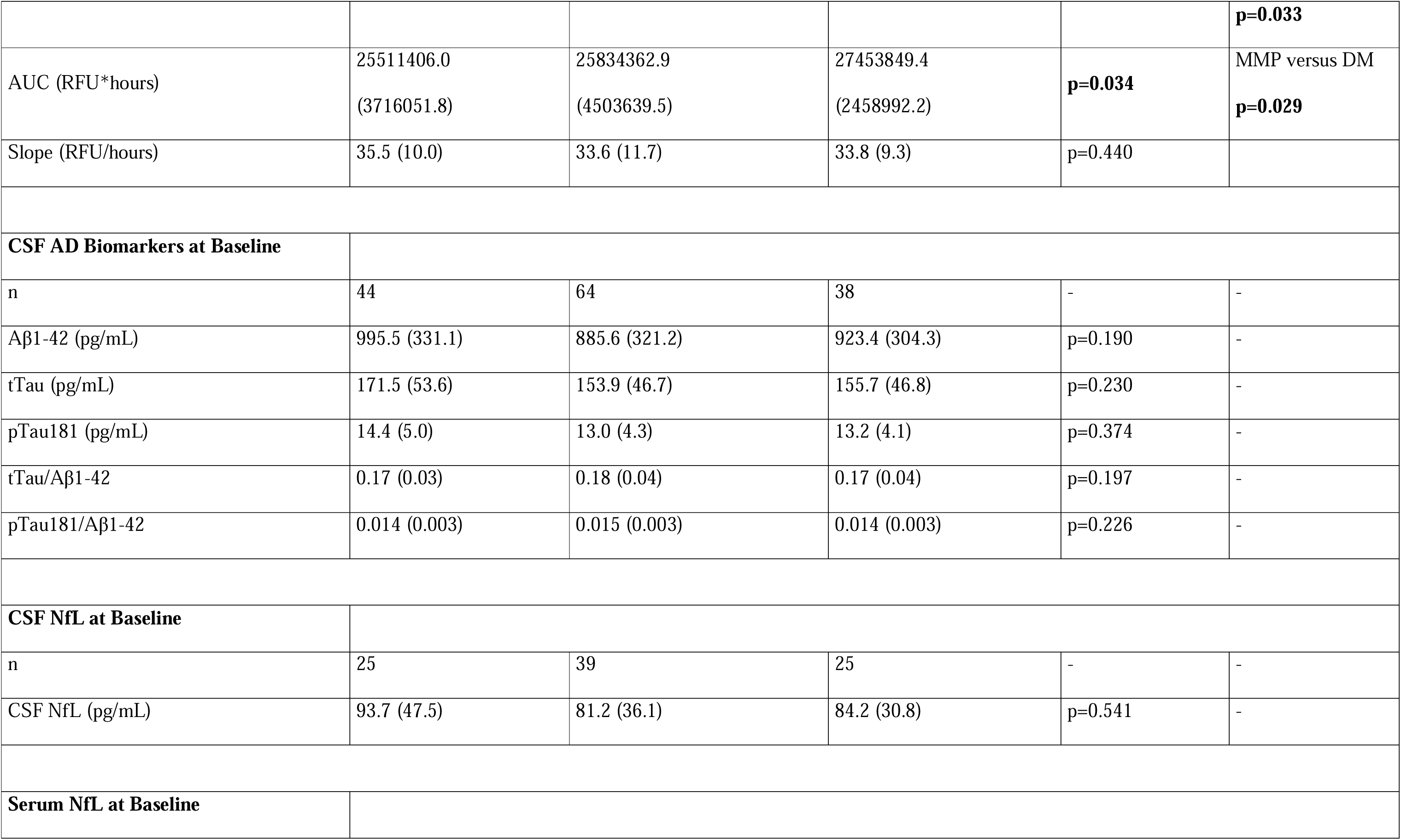
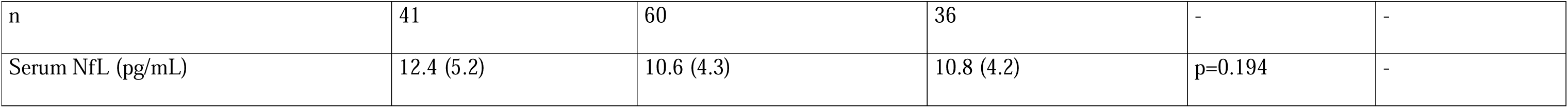
Association between baseline CSF-αSyn-SAA amplification parameters, CSF AD biomarkers, CSF and Serum NfL and Parkinson’s Disease clinical subtype at 10-year follow-up. Values were given in mean (±standard deviation). Statistical significance is marked in bold. Abbreviations: MMP, Mild Motor Predominant; IM, Intermediate; DM, Diffuse Malignant; n, number. Fmax, highest raw fluorescence from each well; T50, time to reach 50% of the Fmax; TTT, time to reach a 5,000 RFU threshold; AUC, area under the curve; AD biomarkers, Alzheimer’s Disease biomarkers; NfL, Neurofilament Light Chain; Aβ1-42, Amyloid Beta Peptide 1-42; tTau, Total Tau; pTau181, Phosphorylated Tau at Serine 181.

**Table 5.** Multinomial Logistic Regression examining the relationship between CSF-αSyn-SAA amplification parameters at baseline and Parkinson’s Disease subtypes at 10-year follow-up. CSF-αSyn-SAA amplification parameters were expressed dichotomously as values below and above the median of each parameter. Only CSF-αSyn-SAA amplification parameters whose mean/distribution significantly differed between 10-year PD subtypes were considered for Multinomial Logistical Regression. CSF-αSyn-SAA amplification parameters were expressed dichotomously as values below and above the median of each parameter. Median of T50: 69.7 hours; Median of TTT: 63.6 hours; Median of AUC: 26909171.8 RFU*hours. Statistical significance is marked in bold. Abbreviations: PD, Parkinson’s Disease; CSF-αSyn-SAA, Alpha-synuclein seed amplification assay on CSF; MMP, Mild Motor Predominant; IM, Intermediate; DM, Diffuse Malignant; T50, time to reach 50% of the Fmax; TTT, time to reach a 5,000 RFU threshold; AUC, area under the curve; OR, odds ratio, CI, confidence interval; χ^2^ (df), Chi-Square (degrees of freedom)

|  |  | **OR** | **95% CI Lower** | **95% CI Upper** | **Sig.** | **Model Fitting Information** |
| --- | --- | --- | --- | --- | --- | --- |

| | | | | | | $\chi^2$ (df) | Sig. |
| --- | --- | --- | --- | --- | --- | --- | --- |
| **T50 below the median** | **DM compared to MMP** | 3.3 | 1.3 | 8.1 | **p=0.010** | 8.187 (2) | **p=0.017** |
|  | **IM compared to MMP** | 2.6 | 1.1 | 5.8 | **p=0.020** |  |  |
| **T50 above the median** | **DM compared to MMP** | 0.3 | 0.1 | 0.7 | **p=0.010** |  |  |
|  | **IM compared to MMP** | 0.4 | 0.2 | 0.8 | **p=0.020** |  |  |
| **TTT below the median** | **DM compared to MMP** | 4.6 | 1.8 | 11.6 | **p=0.001** | 11.662 (2) | **p=0.003** |
|  | **IM compared to MMP** | 2.7 | 1.2 | 6.1 | **p=0.016** |  |  |
| **TTT above the median** | **DM compared to MMP** | 0.2 | 0.1 | 0.5 | **p=0.001** |  |  |
|  | **IM compared to MMP** | 0.4 | 0.2 | 0.8 | **p=0.016** |  |  |
| **AUC below the median** | **DM compared to MMP** | 0.4 | 0.1 | 0.9 | **p=0.031** | 5.047 (2) | p=0.080 |
|  | **IM compared to MMP** | 0.5 | 0.2 | 1.2 | p=0.121 |  |  |
| **AUC above the median** | **DM compared to MMP** | 2.7 | 1.1 | 6.6 | **p=0.031** |  |  |
|  | **IM compared to MMP** | 1.9 | 0.8 | 4.1 | p=0.121 |  |  |

### AD biomarkers, NfL and Clinical Subtypes

Baseline levels of CSF AD biomarkers (including ratios) as well as CSF and Serum NfL did not differ between groups. No statistical difference was noted with either baseline or 10-year follow-up clinical subtypes (Table 3 and 4).

## DISCUSSION

Clinical subtypes of PD can be helpful for patient counseling and prediction of disease progression, but it is uncertain whether they mark distinct clinic-pathological paths. The recent development of CSF-αSyn-SAA as an accurate biomarker of synucleinopathy could provide biological basis for the subtype classifications. In this study, we assessed the 10-year clinical trajectory of MMP, IM and DM subtypes of PD. CSF-αSyn-SAA parameters at baseline were used to characterize each subtype and predict the long-term clinical progression. Markers of axonal degeneration and AD co-pathology were also investigated for a comprehensive analysis.

A half of participants changed subtype during follow-up, mostly shifting towards a more aggressive phenotype. Shorter reaction times (T50 and TTT) and a larger AUC of the CSF-αSyn-SAA amplification was observed in DM subtype compared to IM/MMP subtype. The difference in amplification parameters at baseline was more evident when comparing subtypes established on the 10-year clinical features than when comparing subtypes based on the baseline clinical features.

Therefore, CSF-αSyn-SAA parameters at baseline were predictors of the long-term clinical outcome of PD subjects at 10 years. CSF AD biomarkers, and CSF and serum NfL did not significantly differ between subtypes.

The need to identify PD subtypes aligns with the main goal of precision medicine(20). In the absence of reliable fluid or imaging biomarkers, clinical classifications have been the only feasible strategy so far to this goal. Stratifications based exclusively on PD symptoms however seem to lack consistency over time, which challenges their interpretation(9,10,21,22). We observed a large shift across clinical subtypes after 10-year follow-up as well. Although a bidirectional change was noted, the majority of subjects transitioned to a more malignant phenotype. This supports and extends the similar analyses run over shorter follow-up periods (1 to 5 years)(9,10). Even when considering only the motor symptoms, a tendency to shift from more benign to more severe phenotype over time was shown previously (21–23). These observations highlight the need for a better biological basis for classifying clinical subtypes and disease stages, which appear to shift and overlap partially with progression of the disease.

CSF-αSyn-SAAs includes cyclical process of fragmentation and elongation where αSyn seeds in CSF are amplified incorporating recombinant αSyn into new aggregates, which are detected by fluorescent dyes(12). The speed of the SAA reaction reflects the seed concentration when the assay is performed in vitro using recombinant seeds in artificial buffers (12,14,15). However, human biofluids can introduce factors that affect the amplification process and serial dilution studies show only weak correlation with amplification parameters and clinical and other biomarkers(24). In our analysis, the DM subtype exhibited the shortest T50 and TTT, suggesting a possible higher concentration of seeds. A different speed in the SAA reaction was already observed in PD subjects with versus without cognitive impairment and dysautonomia(15,18,25). Interestingly, the difference in times of reaction was more pronounced across the subtypes based on 10-year follow-up than across the subtypes based on the baseline clinical phenotypes. The MLR confirmed that TTT and T50 below the median at baseline increased the risk of being classified as DM or IM after 10 years by more than 3-4 folds. Moreover, faster SAA reactions were slightly more common in subjects who shifted towards a more malignant subtype at follow-up. Our findings indicate that SAA parameters at the time of diagnosis might have a prognostication value to predict the long-term clinical outcome of PD after 10-years. This is supported by two studies that have addressed this topic to date. The first study reported an association of faster SAA reactions with an earlier development of PD-related dementia(17). The second study identified same biochemical alterations in iRBD subjects who exhibited the highest rate of phenoconversion to PD(19). Significant variability and overlap of amplification parameters were noted across groups in all the above studies including ours, which prevents the use of SAA as a quantitative test at an individual level for now. We found no differences in CSF beta-amyloid, total and phosphorylated tau levels between groups. Beta-amyloid and tau represent the main biological biomarkers to aid AD diagnosis. However, these markers are now also used in PD to define the degree of the AD co-pathology and predict cognitive decline(26). In line with sporadic reports, we were expecting to find reduced CSF Aβ1-42 levels in the DM group, which scored the lowest on the neuropsychological testing as per definition(7). However, subjects enrolled in this study were overall well-preserved in terms of cognitive functions, despite some inter-group differences. The average MOCA score never fell below the threshold for frank dementia. Moreover, the only neuropathological study performed so far did not detect differences in AD co-pathology between MMP, IM and DM subtypes, outlining the need for further investigations(27).

CSF and Serum NfL were also similar in all subtypes. NfL is a marker of large fiber degeneration whose application in PD is limited since their levels are similar to controls, much lower than in MSA, where the rapid neuronal death typically leads to more significant changes in the biomarker concentration (28). The literature reports differences in NfL levels between PD subtypes with distinct prognosis(29–33). Differences between subtypes, however, were consistently very small across all the studies. A single work included MMP, IM and DM subtypes, but the authors used a simplified calculator, limiting a possible comparison between cohorts (34).

Our study has several strengths. First, it is based on a large, well-characterized, international cohort. This enabled the use of a large sample size and long-term follow-up information. Clinical subtypes, MMP, IM and DM, originate from a cluster analysis, which led to unbiased inclusion of a large number of motor and non-symptoms of PD. Although the role of CSF-αSyn-SAA is most robust for diagnosis of synucleinopathies, the current study provides insight into the use of the SAA parameters as a quantitative and predictive test. Among the limitations of the study, data were available only in a limited number of subjects at 10-year follow-up. The amplification parameters are variable and the correlations with clinical subtypes are not robust. The assay data is available only from the baseline, preventing the assessment of their longitudinal change.

In conclusion, our results showed that CSF-αSyn-SAA parameters predict the long-term clinical course of PD better than markers of AD co-pathology or axonal degeneration. These data suggest the use of the assay as a semiquantitative test, but further development of assays that can provide more robust quantitation will be necessary to validate these analyses.

## Acknowledgment

Dr. Kang is supported by NIH (R01 NS131658, U01 NS113851, U01 NS122419, RF1 NS126406, R01 NS133742).

Dr. Riboldi is supported by grants from Michael J Fox Foundation, Parkinson’s Foundation, Department of Defense (PD210038), NIH (R01 NS116006; R01 NS133742), and received a previous research grant from Prevail Therapeutics.

Dr. Grillo is supported by #NEXTGENERATIONEU (NGEU) and funded by the Ministry of University and Research (MUR), National Recovery and Resilience Plan (NRRP), project MNESYS (PE0000006) – A Multiscale integrated approach to the study of the nervous system in health and disease (DN. 1553 11.10.2022). Dr. Grillo is also supported by the Marlene and Paolo Fresco Institute Post-Doctoral Clinical Fellowship.

Dr. Fereshtehnejad is supported by a grant from Parkinson Canada.

Progression Markers Initiative (PPMI) database (www.ppmi-info.org/access-data-specimens/download-data), RRID:SCR_006431. For up-to-date information on the study, visit www.ppmi-info.org.

PPMI – a public-private partnership – is funded by the Michael J. Fox Foundation for Parkinson’s Research and funding partners, including 4D Pharma, Abbvie, AcureX, Allergan, Amathus Therapeutics, Aligning Science Across Parkinson’s, AskBio, Avid Radiopharmaceuticals, BIAL, BioArctic, Biogen, Biohaven, BioLegend, BlueRock Therapeutics, Bristol-Myers Squibb, Calico Labs, Capsida Biotherapeutics, Celgene, Cerevel Therapeutics, Coave Therapeutics, DaCapo Brainscience, Denali, Edmond J. Safra Foundation, Eli Lilly, Gain Therapeutics, GE HealthCare, Genentech, GSK, Golub Capital, Handl Therapeutics, Insitro, Jazz Pharmaceuticals, Johnson & Johnson Innovative Medicine, Lundbeck, Merck, Meso Scale Discovery, Mission Therapeutics, Neurocrine Biosciences, Neuron23, Neuropore, Pfizer, Piramal, Prevail Therapeutics, Roche, Sanofi, Servier, Sun Pharma Advanced Research Company, Takeda, Teva, UCB, Vanqua Bio, Verily, Voyager Therapeutics, the Weston Family Foundation and Yumanity Therapeutics.

## Author Contribution

PG: conception, design, analysis, execution, writing

AP: design, editing of final version of the manuscript

GMR: conception, design, analysis, execution, editing of final version of the manuscript

UJK: conception, design, analysis, execution, editing of final version of the manuscript

SMF: conception, design, analysis, execution, editing of final version of the manuscript

## Statements and Declarations

### Consent for publication

Not applicable

### Funding statement for the preceding 12 months

Dr. Un Kang receives consulting compensation as a SAB member of Amprion, Inc.

Dr. Giulietta Riboldi: nothing to disclose.

Dr. Piergiorgio Grillo: nothing to disclose.

Dr. Antonio Pisani: nothing to disclose.

Dr. Seyed-Mohammad Fereshtehnejad: nothing to disclose.

### Data availability

Data are available from authors upon reasonable request.

